# Assessing Global Covid-19 Cases Data through Compositional Data Analysis(CoDa)

**DOI:** 10.1101/2020.12.17.20248424

**Authors:** Luis P.V. Braga, Dina Feigenbaum

**Affiliations:** Federal University of Rio de Janeiro

**Keywords:** CoDa, LogRatio, Clusterization, Covid-19, JHU

## Abstract

**Background:** Covid-19 cases data pose an enormous challenge to any analysis. The evaluation of such a global pandemic requires matching reports that follow different procedures and even overcoming some countries’ censorship that restricts publications.

**Methods:** This work proposes a methodology that could assist future studies. Compositional Data Analysis (CoDa) is proposed as the proper approach as Covid-19 cases data is compositional in nature. Under this methodology, for each country three attributes were selected: cumulative number of deaths (D); cumulative number of recovered patients(R); present number of patients (A).

**Results:** After the operation called closure, with c=1, a ternary diagram and Log-Ratio plots, as well as, compositional statistics are presented. Cluster analysis is then applied, splitting the countries into discrete groups.

**Conclusions:** This methodology can also be applied to other data sets such as countries, cities, provinces or districts in order to help authorities and governmental agencies to improve their actions to fight against a pandemic.

## 1. Introduction

Among several difficulties to analyze the state of the pandemic is the complexity of testing logistics and getting the right results. There are also the political restrictions, as in many countries censorship restricts the publication of surveys and reports. Besides that, if you wish to evaluate globally the pandemic census you have to match different reports from many countries that have their own methodology. The Johns Hopkins University(JHU) assumed the huge task of providing an account of the pandemic worldwide^1^. JHU data are used in this work as it is described in the dataset section.

The purpose of this work is to explore the relationship between the proportions of the attributes by applying Compositional Data Analysis (CoDa)^2^. Three attributes were selected for each country: cumulative number of deaths (D); cumulative number of recovered patients(R); present number of patients (A). After the operation called closure, with *c*=1 (acomp scale) a ternary diagram and Log-Ratio plots, as well as, basic compositional statistics were obtained. Cluster analysis was then applied, splitting the countries into groups. The results must be understood as descriptive epidemiologic assumptions, even though some associate patterns are also suggested^3^, they have an interpretation coherent with the spread of the pandemic.

## 2. Methodology

The Compositional Data contains information about relative magnitudes. Dependency among variables of a composition can be examined in real space by analyzing the covariance structure of the log-ratios. The compositional approach consists of a change of representation of the original sample space, the simplex S^D^, onto a new sample space, namely real space D–1.

Components are the individual parts x_i_ of the composition ***x***. The sum over the amount of all components ***c*** is called total, **c** usually is 1:

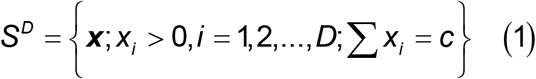

The closure of a set is defined by:

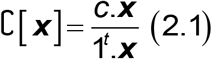

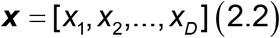

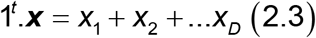

It is proved that the simplex, with the operations of perturbation, powering and Aitchinson inner product, has a (D-1)-dimensional Euclidean vector space structure. So through an isometric transformation virtually anything could be translated from real vectors to compositions and vice versa^4^, such as the Center Log-Ratio and the Isometric Log-Ratio Transformations.

### Centered Log-Ratio Transformation (clr)

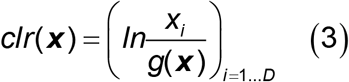

g is the geometric mean of the components.

### Isometric Log-Ratio Transformation (ilr)

It is an isometric transformation between the simplex S^D^ and the (D-1) dimensional real space. A set of (D-1) orthornormal directions is a base, called balance base. An ilr transformation of a composition is the projection onto this base^5^.

### Descriptive Analysis of Compositional Data

The arithmetic mean and the variance or standard deviation of individual parts do not fit as the value of central tendency and measure of dispersion because the parts of a composition are linked to each other and they are multivariate by nature. Taking this into account, it follows that the center (cen) or compositional mean of a data set **X** with N observations and D parts is a composition, i.e., the closed geometric mean. It is defined as:

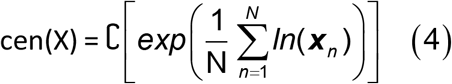

The variation matrix T describes the dispersion in a compositional set. It has D^2^ components, each defined as:

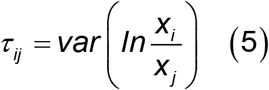

The smaller the matrix variation element is, the better the proportionality between the two components.

### Clusterization

The grouping of samples presupposes the definition of two rules - the distance formula between two samples and an agglomeration criterion. This subject is well known in the Euclidean case, where several distances are possible: Euclidean, Manhattan, Mahalanobis, Minmax. Most distance measures for multivariate datasets can be generalized to composition formalism^6^.

The Aitchison’s distance formula is:

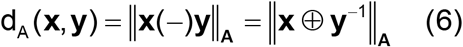

In other cases, it is necessary to adapt the formulas by using the Centered Log-Ratio transformation (crl) as it is done for the Euclidean distance in this work or the Isometric Log-Ratio transformation (ilr).

Once the distance formula has been defined, the next step should be the definition of the crowding criteria. Among others, in addition to Ward’s criterion^7^ there are the simple, medium and complete binding criteria.

In the Ward’s method^7^, for a given group, the distance of each external object to the group average is calculated and the object that causes the smallest increase in the sum of distances is incorporated. The connection criteria allow to calculate the distance between elements and, also, between groups of elements. The simple option considers that the distance between two groups (unitary or not) is the shortest distance possible between an element in one group and an element in another one. The average option calculates the average distance for all possible pairs. Furthemore, the complete option considers that the distance between groups corresponds to the greatest possible distance between an element from one group and an element from another one.

There are several packages that implement the CoDa theory. The software “Compositions” ^8^ in “R” ^9^ was the one adopted.

### Data

In this study the data sources are WHO, CDC, ECDC, NHC, DXY, 1point3acres, Worldometers.info, BNO, the COVID Tracking Project (testing and hospitalizations), State and National Government Health Departments, and local media reports. A layer in the package ArcGis^10^ was created and maintained by the Center for Systems Science and Engineering (CSSE) at the Johns Hopkins University (CSSE 2020). This feature layer is supported by ESRI Living Atlas team, JHU APL and JHU Data Services. This layer is opened to the public and free to share. The cases dataset was downloaded from that repository on the 6^th^ of September, 2020, and includes the following attributes: Country Name, Deaths(D), Recovered(R) and Active(A) patients. Note that the second and the third are cumulative figures until that day and the last one is the value available on that day. The raw cases data are displayed in Appendix I and the closure (acomp scale) in Appendix II, both are available at http://dx.doi.org/10.17632/wt7nd5jv6s.1

For each frequency (Deaths, Recovered, Active) proportions of the population were calculated in the acomp scale, entries with null values were converted to Below Detection Limits (BDL). The original attributes will now be named in the acomp scale:

P_D_: cumulative relative frequency of deaths (8)

P_R_: cumulative relative frequency of recovered (9)

P_A_: relative frequency of active^*^ (10)

P_c_: frequency (cumulative and 6^th^ of September) of confirmed^**^ (11)

*People still being treated

**People that caught the Covid-19

[P_D_, P_R_, P_A_] shows the inner rapports between the confirmed categories.

## 3. Results

The results are divided into two categories: compositional descriptive and compositional associative.

### Compositional Exploratory Analysis of the Subcomposition

[P_D_, P_R_, P_A_]

In this case the closure is performed over the three components and the values represent relative proportions among them. The null counts are treated as “Below Detection Limit”, therefore to input the data in the package “Compositions” they must be converted to −1, these cases are depicted as red lines in the ternary diagrams below.

**Table 1.**
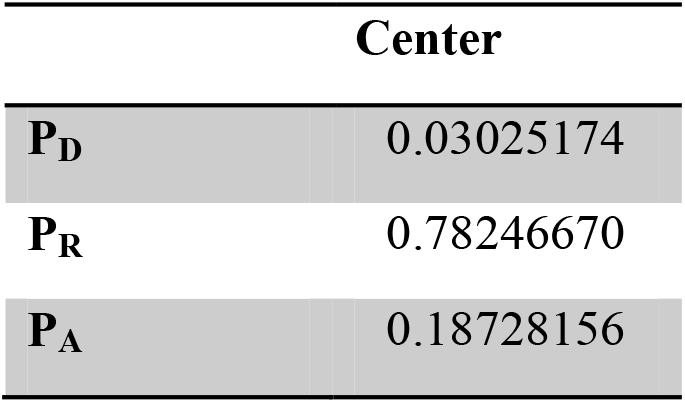
Center

This result shows that the P_R_ is the main overall feature.

**Table 2.** Variation Matrix

| | $P_D$ | $P_R$ | $P_A$ |
| --- | --- | --- | --- |
| $P_D$ | 0.000000 | **1.380129** | 2.028660 |
| $P_R$ | **1.380129** | 0.000000 | 2.386253 |
| $P_A$ | 2.028660 | 2.386253 | 0.000000 |

The components P_D_ and P_R_ are more proportional than P_D_ and P_A_ or P_R_ and P_A_, formula (5), what is understandable because the Active patients will turn into the category of either Recovered or Dead, A => R or D.

### Plots of samples against a Log-ratio

The following analysis will show the rapport between the proportions in the composition [PD, PR, PA] through the signal of the Log-ratio between them. The results will be displayed in non-centralized ternary diagrams in order to highlight both the relationship between countries and the attributes. In the diagram, the closer a country is to a vertex, the greater the proportion of the attribute associated with it in relation to the other attributes.

In this figure the following countries were not depicted: Buthan, Cambodia, Djibouti, Dominica, Eritrea, Grenada, Holy See, Laos, Mongolia, Saint Kitts and Nevis, Saint Lucia, Saint Vincent and Grenadine, Serbia, Seychelles, Sweden and Timor Leste. The reason is because they presented null values either on Deaths or on Active patients, therefore P_D_, P_R_ or P_A_=0.

### Log-Ratio Scatterplot P_D_/P_R_

This plot, fig. 1a and b, shows the lack of effectiveness of the country to save lives against the recovery of patients. At the date of the evaluation, The Netherlands and The United Kingdom had the greatest proportion of deaths over recoveries, as well as Qatar.

**Fig. 1.**
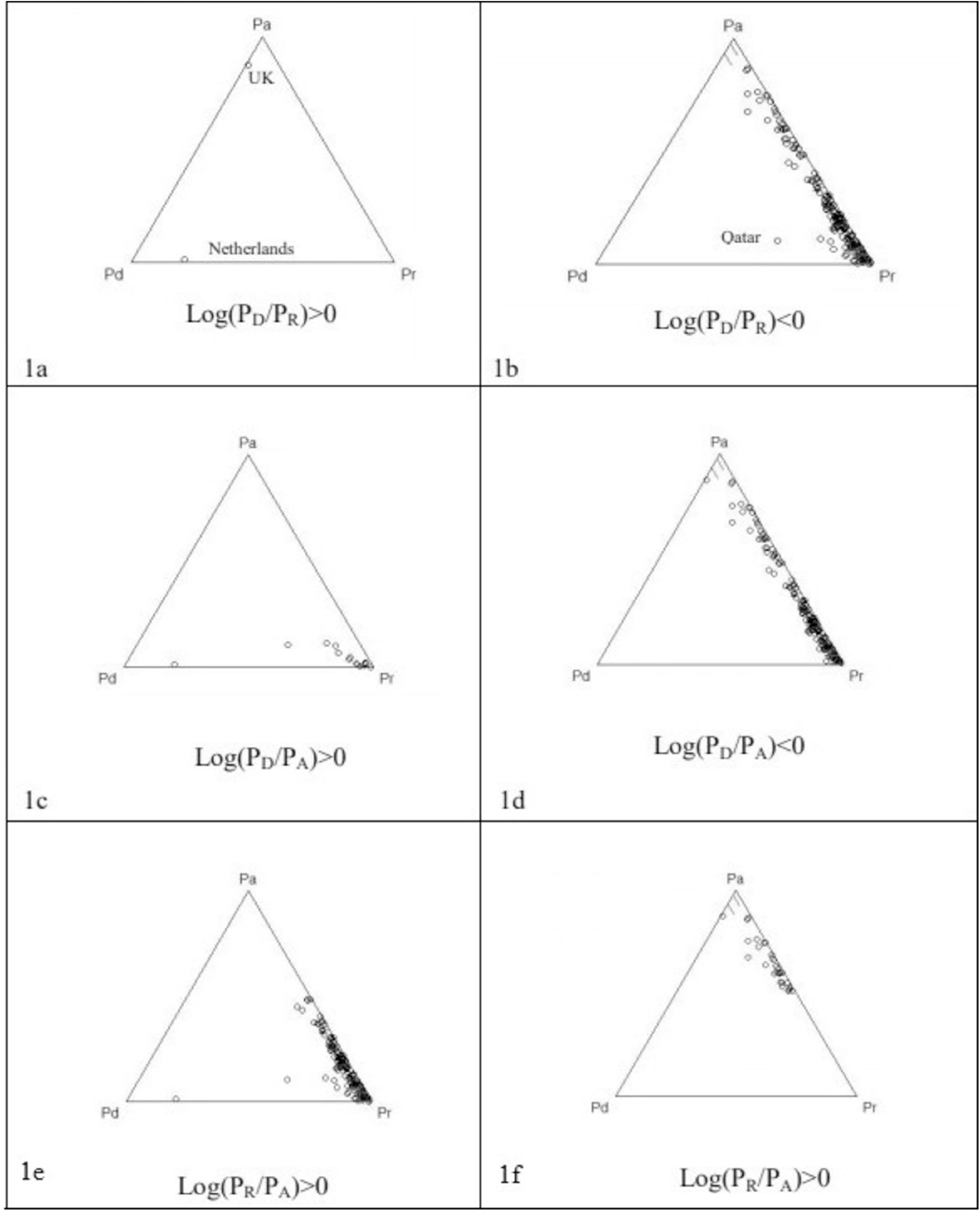
Plots of samples against a Log-ratio

**Fig. 2.**
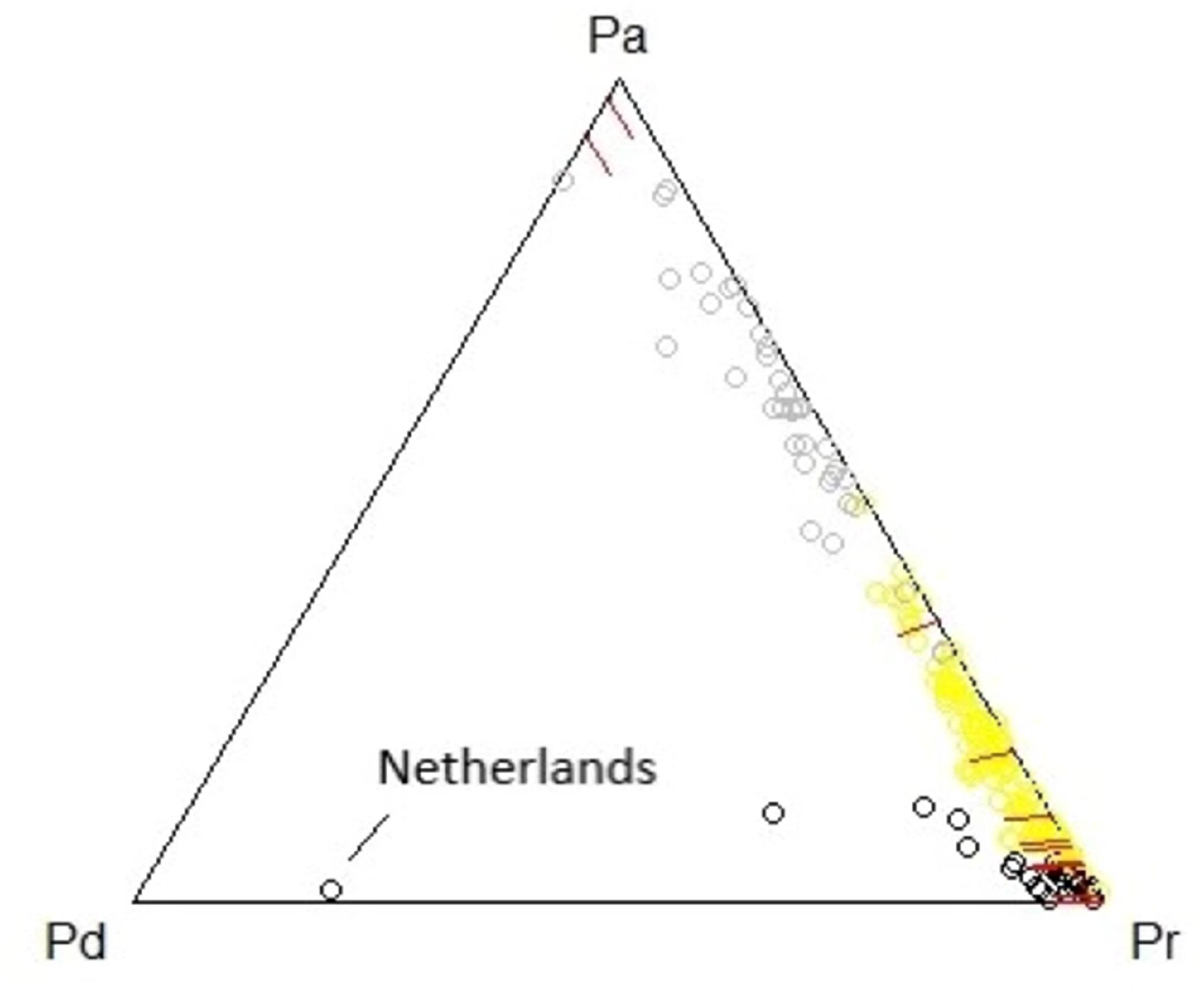
Groups in a ternary diagram

### Log-Ratio Scatterplot P_D_/P_A_

This plot, fig. 1c and d, shows the country loss of lives among infected people. At the date of the evaluation. In Netherlands the past cases are pushing the log ratio up, the country had a total of 6277 deaths against 116 active patients.

### Log-Ratio Scatterplot P_R_/P_A_

This plot, fig1.e1e and f shows the effectiven_1_e_f_ ss of the country ability to recover lives among infected people. At the date of the evaluation. The United Kingdom presented a very low proportion of recovering and the largest proportions of active patients.

The bisectors in the ternary diagrams determine the samples order relations. For instance:

P_D_>P_R_ => samples belonging to the bisector defined by P_A_ and the edge

P_A_P_D_ P_D_>P_A_ => samples belonging to the bisector defined by P_R_ and the edge

P_R_P_D_ P_R_>P_A_ => samples belonging to the bisector defined by P_D_ and the edge P_D_P_R_

### Clusterization

The final step of the analysis provided an insight on how the groups of countries are formed. From a pure mathematical criterion it was possible to uncover order relations between the countries. An hierarchical clusterization algorithm following the Ward criterion was applied to the subcomposition [P_D_,P_R_, P_A_] resulting in 3 different groups for height=10 in the hierarchical tree. The Euclidean distance was calculated after a clr transformation of the acomp transformed data was done.

The groups are shown in different colors. The table 3 discriminates each country by group. The interpretation of the clustering is done in terms of the order relations between the different categories.

The gray group is closer to Pa, the yellow closer to Pr and the black between Pd and Pr. The three groups reproduce the categories seen in the ternary diagram with the additional information on the log-ratios. The gray group has lower proportions of dead and recovered. The yellow group has intermediate levels of these proportions and the black group is composed of dominant proportions of dead or recovered.

Table 3

Countries per group. A full detailed table is available in:

http://dx.doi.org/10.17632/wt7nd5jv6s.1

### Group gray

Angola, Bahamas, Bangaladesh, Belgium, Belize, Bhutan, Botswana, Burma, Central African Republic, Congo (Brazaville), Costa Rica, Czequia, Ethiopia, France, Gambia, Greece, Grenada, Guinea-Bissau, Holy-See, Honduras, Hungary, Jamaica, Kenya, Laos, Lebanon, Lesotho, Lybia, Namibia, Papua New Gune, Paraguay, Romania, Saint Kitts Nevis, Saint Lucia, Saint Vincent Grenadines, Spain, Sudan, Sweden, Syria, Tanzania, Trinidad Tobago, Tunisia, Uganda, Ukraine, United States

Countries in this group have Deaths < Recovered < Active, see Appendix I for more details. It implies also that P_D_ < P_R_ < P_A_. It is a group **Active dominant and Recovery subdominant**.

### Group yellow

Afghanistan, Albania, Algeria, Andorra, Argentina, Armenia, Australia, Austria, Azerbaijan, Bahrain, Barbados, Benin, Bolivia, Bosnia Herzegovina, Brazil, Bulgaria, BurkinaFaso, Burundi, Cabo Verde, Colombia, Comoros, Cote de Ivoire, Croatia, Cuba, Cyprus, Denmark, Djibouti, Dominican Republic, Ecuadro, Egypt, El Salvador, Equatorial Guinea, Estonia, Eswatini, Fiji, Finland, Gabon, Georgia, Germany, Guatemala, Guinea, Guyana, Haiti, Iceland, India, Indonesia, Iran, Iraq, Ireland, Israel, Japan, Jordan, Kazakhstan, Korea South, Kosovo, Kuwait, Kyrgyzstan, Latvia, Liechtenstein, Lithuania, Luxembourg, Madagascar, Malawi, Maldives, Mali, Malta, Moldavia, Monaco, Montenegro, Morocco, Mozambique, Nepal, New Zealand, Nicaragua, Nigeria, North Macedonia, Norway, Oman, Panama, Peru, Philippines, Poland, Protugal, Qatar, Russia, Rwanda, Saudi Arabia, Senegal, Sierra Leone, Singapore, Slovakia, Slovenia, Somalia, South Africa,Sri Lanka, Suriname, Switzerland, Tajikistan, Togo, Turkey, United Arab Emirates, Uruguay, Uzbekistan, Venezuela, West Bank Gaza, Zambia, Zimbabwe

Countries in this group have Deaths < Recovered; Deaths < Active and Recovered > Active, see Appendix I for more details. It implies also that P_D_ < P_R_; P_D_ < P_A_ and P_R_ > P_A_. It is a group **Recovery Dominant and Active subdominant**.

### Group black

Antigua-Barbuda, Belarus, Brunei, Cambodia, Cameron, Canada, Chad, Chile, China, Congo (Kinghasa), Dominica, Eritrea, Ghana, Italy, Liberia, Malaysia, Mauritania, Mauritius, Mexico, Mongolia, Netherlands, Niger, Pakistan, San Marino, Sao Tome Principe, Seychelles, Taiwan, Thailand, Timor Leste, Vietnam, Western Sahara, Yemen

Countries in this group have Deaths > Active; Recovery > Deaths and Recovery > Active, see Appendix I for more details. It implies also that P_D_ > P_A_; P_R_ > P_D_ and P_R_ > P_A_. It is a group **Recovery Dominant and Deaths subdominant** or **Active Dominant and Death subdominant**.

**T**he lists in the table above just show vis a vis the past incidences, the potential stages of the pandemic.

## 5. Conclusion

The Composition Data Analysis (CoDa) theory is the natural choice for uncovering the hidden aspects of the pandemic cases. Developed and wealth countries surprised the world as their populations were vastly touched by the virus with large numbers of deaths and sick people. However, there are restrictions with the data as they come from different sources and, as in the Spanish fever, frequently censored^11^. Spite of those drawbacks, the CoDa was able to unveil a few trends in the pandemic.

The levels of the proportions of deaths, recovered and active cases revealed unexpected outliers, as well as a common behaviour among countries. The ternary diagram, the log ratio plots and the clusterization suggested three categories of countries: (black) those recovery dominant, but death subdominant, as on the 6^th^ of september had more deaths than active; (yellow) a very big group which is recovery dominant and at last, the (gray) group which is recovery dominant and deaths subdominant or active dominant and death subdominant. None group is the good or the bad group, they just show different characteristics like what has been observed in the Log-Ratio plots. The method can be applied to provinces in a country, or counties in a state in order to support public action planning

This study is a snapshot of the pandemic on the 6^th^ of September, it does not make any inference about the rate, or the aftermath of the first wave. The purpose was to show the ability of the CoDa theory to unveil the proportions between the three different types of cases: Deaths, Recoveries and Active. A dynamical study seems to be the natural sequence of this approach, as well as the production of inference models. This methodology can be applied to further data sets such as countries, cities, provinces or districts in order to help authorities and governmental agencies to improve their actions to fight the pandemic^12^.

### CRediT authorship contribution statement

Luis P. V. Braga: Conceptualization, Methodology, Software, Data Curation, Writing - Original Draft.

Dina Feigenbaun: Conceptualization, Methodology, Writing - Review & Editing

## Data Availability

In this study the data sources are WHO, CDC, ECDC, NHC, DXY,1point3acres, Worldometers.info, BNO, the COVID Tracking Project (testing and hospitalizations), State and National Government Health Departments, and local media reports. A layer in the package ArcGis10 was created and maintained by the Center for Systems Science and Engineering (CSSE) at the Johns Hopkins University (CSSE
2020). This feature layer is supported by ESRI Living Atlas team, JHU APL and JHU
Data Services. This layer is opened to the public and free to share. The cases dataset was downloaded from that repository on the 6th of September, 2020, and includes the following attributes: Country Name, Deaths(D), Recovered(R) and Active(A) patients.
Note that the second and the third are cumulative figures until that day and the last one
is the value available on that day. The raw cases data are displayed in Appendix I and the closure (acomp scale) in Appendix II, both are available at
http://dx.doi.org/10.17632/wt7nd5jv6s.1

## References

1. Dong E, Hongru D, Gardner L An interactive web-based dashboard to track COVID-19 in real time. The Lancet Infectious Disease, 2020 Volume 20 Number 5 p511-628, e79-e115. https://www.thelancet.com/action/showPdf?pii=S1473-3099%2820%2930120-1

2. Filmoser P, Hron K, Templ M. Compositional Data as a Methodological Concept., In: Applied Compositional Data Analysis. Springer Series in Statistics. Springer, Cham 2018; https://doi.org/10.1007/978-3-319-96422-5_1

3. Wasserthel-Smoljers S. Biostatistics & epidemiology: a primer for health professionals. New York: Springer, 1990. ISBN: 978-1-4757-3887-2 (eBook)

4. Pawlowsky-Glahn V, Egozcue J, Tolosana-Delgado R. R Modeling and Analysis of Compositional Data. 1^a^ Ed. West Sussex: Wiley, 2015. ISBN: 9781118443064

5. Egozcue J, Pawlowsky-Glahn V, Mateu-Figueras G, Barceló-Vidal C. Isometric logratio transformations for compositional data analysis. Mathematical Geology, 2003 Volume 35, Number 3 p279-300.

6. Boogaart K G, Tolosana-Delgado R. Analyzing Compositional Data with R. Heidelberg: Springer, 2013. ISBN 978-3-642-36808-0

7. Murtag F, Legendre P. Ward’s Hierarchical Agglomerative Clustering Method: Which Algorithms Implement Ward’s Criterion? Journal of Classification, 2014 Volume 31, p274-295.

8. Boogaart K G, Tolosana-Delgado R. Compositional data analysis with ‘R’ and the package ‘compositions’. Geological Society, 2006. Special Publications 264, p119-127.

9. R CORE TEAM. R: A language and environment for statistical computing. R Foundation for Statistical Computing, 2020. Vienna, Austria; https://www.r-project.org/

10. ARCGIS: The mapping and analytics platform. https://www.esri.com/en-us/arcgis/about-arcgis/overview

112. Barry J. How the Horrific 1918 Flu Spread Across America. Smithsonian Magazine, 2017 November. https://www.smithsonianmag.com/history/journal-plague-year-180965222/#.XmRY91BulqA.linkedin

12. McKay S, Boyce M, Chuh-Shin S, Tsai F J, Katz R. An Evaluation Tool for National-Level Pandemic Influenza Planning. World Medical & Health Policy, 2019, Volume 11, Number 2, p127-133.

